# Resilient T cell responses to B.1.1.529 (Omicron) SARS-CoV-2 variant

**DOI:** 10.1101/2022.01.16.22269361

**Authors:** Mladen Jergovic, Christopher P. Coplen, Jennifer L. Uhrlaub, Shawn C. Beitel, Jefferey L. Burgess, Karen Lutrick, Katherine D. Ellingson, Makiko Watanabe, Janko Nikolich-Žugich

## Abstract

Emergence of the SARS-CoV-2 variant-of-concern (VOC) B.1.1.529 (Omicron) in late 2021 has raised alarm among scientific and health care communities due to a surprisingly large number of mutations in its spike protein. Public health surveillance indicates that the Omicron variant is significantly more contagious than the previously dominant VOC, B.1.617.2 (Delta). Several early reports demonstrated that Omicron exhibits a higher degree (∼10-30-fold) of escape from antibody neutralization compared to earlier lineage variants. Therefore, it is critical to determine how well the second line of adaptive immunity, T cell memory, performs against Omicron in people following COVID-19 infection and/or vaccination. To that purpose, we analyzed a cohort (n=345 subjects) of two-or three-dose messenger RNA (mRNA) vaccine recipients and COVID-19 post infection subjects (including those receiving 2 doses of mRNA vaccine after infection), recruited to the CDC-sponsored AZ HEROES research study, alongside 32 pre-pandemic control samples. We report that T cell responses against Omicron spike peptides were largely preserved in all cohorts with established immune memory. IFN-γ producing T cell responses remained equivalent to the response against the ancestral strain (WA1/2020), with some (<20%) loss in IL-2 single-or IL-2+IFNγ+ poly-functional responses. Three-dose vaccinated participants had similar responses to Omicron relative to convalescent or convalescent plus two-dose vaccinated groups and exhibited responses significantly higher than those receiving two mRNA vaccine doses. These results provide further evidence that a three-dose vaccine regimen benefits the induction of optimal functional T cell immune memory.

## Introduction

The B.1.1.529 (Omicron) SARS-CoV-2 variant likely originated in Botswana and was first reported to the World Health Organization (WHO) by the South African government on November 24^th^, 2021^1^. Several days later, the WHO and the United States designated Omicron as a variant of concern (VOC). Omicron has accumulated a surprisingly large number of mutations compared to the original Wuhan strain; a total of 60 out of which 50 were nonsynonymus^2^. 32 of those mutations reside in the Spike (S) protein, which is the sole target of the majority of the currently approved SARS-CoV-2 vaccines and immunoglobulin therapies. The large number of mutations raised concerns about the potential for increased transmissibility, escape from vaccine protection, and potential pathogenicity. Although peer-reviewed epidemiological and clinical studies remain scarce, considerable evidence points to increased transmissibility of Omicron as compared to previous VOCs, including the highly transmissible B.1.617.2 (Delta) variant. A cohort analysis of household transmission with >72 thousand index cases in England showed that two-fold more Omicron index cases gave rise to a secondary household case in comparison to Delta^3^. Omicron is now the dominant SARS-CoV-2 variant in many parts of the world^4,5^, with a recent study from the University of Hong Kong reporting that the Omicron variant multiplies about 70 times faster in human respiratory tract tissues as compared to Delta^6^. Modeling studies further showed that mutations in the receptor binding domain (RBD) result in stronger binding of Omicron S protein to human ACE2 receptor^7^. Early studies demonstrated a substantial reduction in antibody neutralizing capacity against Omicron, and *in vitro* findings using authentic SARS-CoV-2 viruses indicated that compared to Delta the neutralization efficacy of vaccine-elicited sera against Omicron was severely reduced^8^.

This was independently confirmed by the findings of Rössler et al. which demonstrated that antibody neutralization capacity against Omicron was maintained best in sera from individuals who experienced infection and were subsequently vaccinated^9^.

Of interest, the spread of Omicron in South Africa has been followed by a decrease in hospitalizations^10^ and death rates^11^. Latest reports from UK also showed lower hospitalization rates and mortality over the period when Omicron became the dominant VOC^3^. Thus, the current consensus is forming that disease severity may be reduced with the Omicron variant as compared to Delta. Given the large degree of escape from antibody response, T-cell mediated immunity could be essential to prevent Omicron-induced severe COVID-19. Indeed, in studies with the original, Wuhan strain, depletion of T cells in convalescent macaques resulted in impaired immunity against rechallenge with SARS-CoV-2, suggesting a significant role for T cells in the context of subprotective antibody titres^12^. Human clinical studies also demonstrated a link between SARS-CoV-2 T cell responses and reduced disease severity^13,14^. Thus, it is paramount to determine how well T cell immunity against Omicron is preserved in vaccinees and COVID-19 convalescents. We and others have shown that T cell immunity against Delta, Gamma and other variants was largely preserved in mRNA vaccine recipients^15,16^. However, earlier variants contained drastically lower numbers of mutations in the spike protein.

To assess cross-reactive T cell immune memory to the Omicron variant we have analyzed a large cohort of participants recruited into the CDC-sponsored AZ HEROES research study^17^. Participants in this cohort included subjects sampled post SARS-CoV-2 infection; fully vaccinated subjects (2 doses of mRNA vaccine); subjects post infection receiving 2 doses of mRNA vaccine; fully vaccinated and boosted (three doses of mRNA vaccine) participants; as well as pre-pandemic controls. We have measured polyfunctional T cell responses in peripheral blood mononuclear cells (PBMCs) of 377 participants to overlapping peptide pools corresponding to SARS-CoV-2 S proteins of USA-WA1/2020 (ancestral strain) and the Omicron variant. We report that T cell responses to spike protein of the Omicron variant are largely preserved in both vaccinated and post infected subjects. We found no waning of T cell IFN-γ responses upon stimulation with Omicron spike peptide pools compared to original strain (USA-WA1/2020), with a slight reduction (<20%) in IL-2-producing and polyfunctional IFN-γ+IL-2+ double-positive cells. Additionally, we report that T cell responses in participants receiving two doses of mRNA vaccines were lower than post infection, post infection + 2 vaccine doses and in those receiving three vaccine doses (two + booster), suggesting that three doses of mRNA should be considered as an optimal, full vaccination regimen to induce robust T cell immunity.

## Materials and methods

### Study participants

This study was approved by the University of Arizona IRB (protocol #2102460536 and #1410545697) and the Oregon Health and Science University IRB (protocol #00003007, pre-pandemic controls) and was conducted in accordance with all federal state and local regulations and guidelines. The cohort included 215 recipients of two doses of mRNA SARS-CoV-2 vaccines, 25 recipients of three doses of mRNA SARS-CoV-2 vaccines, 60 SARS-CoV-2 positive individuals (confirmed positive by clinical PCR test), 45 SARS-CoV-2 positive individuals vaccinated post infection and 32 healthy community-dwelling individuals recruited in Tucson, Arizona and Portland, Oregon prior to December 2019. All subjects > 55 years of age were considered older adults and subjects younger than 50 were considered adult. We did not include subjects aged 50-55. Blood was drawn into BD Vacutainer Blood Collection Tubes with Sodium Heparin (BD Bioscience, Franklin Lakes, NJ). Peripheral blood mononuclear cells were separated by ficoll gradient separation and cryopreserved in fetal calf serum + 10% DMSO. PBMC samples were stored in liquid N_2_ at the University of Arizona biobank.

### FLUORISpot assays

Cryopreserved PBMC (5 × 10^6^/sample) were thawed in prewarmed RPMI-1640 media supplemented with L-glutamine (Lonza, Basel, Switzerland) + 10% FCS. Thawed PBMCS were rested for 3-4h at 37 _C in X-VIVO™ 15 Serum-free Hematopoietic Cell Medium (Lonza) supplemented with 5% human AB serum. Cells were then stimulated with ∼1 nmol of peptide pool corresponding to SARS-CoV-2 spike (S) of US-WA (wild-type) or Omicron (B.1.1.529) variant (16-mer peptide pools, overlapping by 10 amino acids, purchased from 21st century Biochemicals Inc.). Cell suspensions were transferred to pre-coated Human IFN-γ, TNF-α, IL-2, Granzyme-B (GrB) FLUORISpot kits (Mabtech, Inc.) and developed after 48h according to manufacturer instructions. Spots were imaged and counted using a Mabtech Iris Fluorispot reader (Mabtech). Samples with low post thaw viability (<60%) were excluded from the analysis, this included 5 samples from recipients of two doses of mRNA SARS-CoV-2 vaccines, 1 sample from recipients of three doses of mRNA SARS-CoV-2 vaccines, 6 samples from SARS-CoV-2 positive individuals (confirmed positive by clinical PCR test), and 6 samples form SARS-CoV-2 positive individuals vaccinated post infection.

### Flow cytometry

Cryopreserved PBMC (5 × 10^6^/sample) were thawed in prewarmed RPMI-1640 with L-glutamine (Lonza, Basel, Switzerland) + 10% FCS. Thawed PBMCS were rested overnight at 37 _C in X-VIVO™ 15 Serum-free Hematopoietic Cell Medium (Lonza) supplemented with 5% human Ab serum. Cells were stained with surface antibodies in PBS (Lonza) + 2% FCS with a total of 11 antibodies (full list of antibodies and fluorochromes in Suppl. Table 1), stained with the live dead fixable blue dye (Thermofisher) and then fixed and permeabilized using the FoxP3 Fix/Perm kit (eBioscience, San Diego, CA). Samples were acquired using a Cytek Aurora cytometer (Cytek, Fremont, CA) and analyzed by FlowJo software (Tree Star, Ashland, OR). Dead cells and doublets were excluded prior to analysis.

### Statistical analysis

Graph Pad Prism v9 was used for statistical analysis. Upon inspection of data distribution by Shapiro-Wilks normality test, differences between paired samples treated with different peptide pools were calculated by two-tailed Wilcoxon rank test. Differences between the subject groups were calculated either by Mann Whitney U-test, Kruskal Wallis test with Dunn’s post hoc correction and. For all statistical differences *p<0.05, **p<0.01, ***p<0.001. ****p<0.0001.

## Results and Discussion

A total of 377 participants were included in this study: (i) 215 received two doses of SARS-CoV-2 mRNA vaccines (2X VAX); (ii) 60 were SARS-CoV-2 post infection samples with no additional vaccination (PI); (iii) 45 were vaccinated following SARS-CoV-2 infection (PI + VAX); (iv) 25 received three doses of mRNA vaccines (3X VAX); and (v) 32 participants presumed immunologically naïve to SARS-CoV-2, recruited before 09/2019 (participant demographics in **Table 1**). We have analyzed T cell responses of the participants to the spike protein of ancestral SARS-CoV-2 (USA-WA1/2020) and the newly emerged B.1.1.529 Omicron variant by stimulating PBMCs with overlapping peptide pools spanning the entire spike protein and measuring production of T cell effector cytokines IFN-γ, IL-2 and Granzyme B (GrB) by FLUORISpot. For each sample, the number of spike-specific spot forming units (SFU) of each cytokine was calculated by subtracting the SFU number measured in the unstimulated wells from the SFU number of experimental, spike peptide stimulated wells (representative FLUORISpot images in **Figure 1A**). Spike peptide pool induced a 5-fold increase in IFN-γ SFU in immunized post pandemic participants while no effect was evident in the pre-pandemic samples (**Suppl. Figure 1A**). Pre-pandemic samples overall showed little crossreactive responses to SARS-CoV-2 peptide pools. This limited cross reactivity was slightly higher against the WA1/2020 peptide pool compared to Omicron, as evidenced by a higher number of IFN-γ SFU (**Suppl. Figure 1B**). When comparing samples from all immunized participants, the number of IFN-γ SFU between Omicron and WA1/2020 peptide stimulated wells was equal (**Figure 1B**, left panel). However, the number of IL-2 SFU was reduced (Figure 1B, middle panel) from an average of 167 with WA1/2020 to 139 with the Omicron peptide pool, representing a 16.8% reduction. There was no difference in GrB SFU between the peptide pools (**Figure 1B**, right panel). Next, we analyzed the number of polyfunctional (double or triple cytokine positive) T cells. We detected a decrease in IFN-γ IL-2 double positive cells, but not IFN-γ GrB double positive cells (**Figure 1C**), while triple positive (IFN-γ, IL-2, GrB) were also decreased with Omicron peptide pool stimulation. Based on this large sample (N=345 exposed/immunized participants) we can conclude that the decrease in T cell immunity to mutated Omicron spike peptide is minimal and that functional responses are largely preserved.

**Table 1.** Demographics of study participants. NA – not applicable.

|  | Pre-pandemic<br>CTRLS | SARS-CoV-2<br>PI | PI + VAX | 2X VAX | 3X VAX |
| --- | --- | --- | --- | --- | --- |
| **Total number of subjects** | 32 | 60 | 45 | 215 | 25 |
| **Gender (n)** |  |  |  |  |  |
| **Male** | 11 | 28 | 22 | 70 | 8 |
| **Female** | 21 | 32 | 23 | 145 | 17 |
| **Age, (mean ± SD)** | 51.5 ± 24.4 | 50.8 ± 13.3 | 50.7 ± 13.2 | 50.4 ± 11.7 | 58.1 ± 22.1 |
| **Vaccine Manufacturer (n)** |  |  |  |  |  |
| **Pfizer** | NA | NA | 23 | 133 | 12 |
| **Moderna** | NA | NA | 22 | 82 | 11 |
| **Days post VAX (mean ± SD)** | NA | NA | 125.0 ± 80.4 | 169.8 ± 75.3 | 33.3 ± 24.7 |
| **Days post COVID onset**<br>**(mean ± SD)** | NA | 182.6 ± 102.6 | 264.9 ± 107.1 | NA | NA |

**Figure 1.**
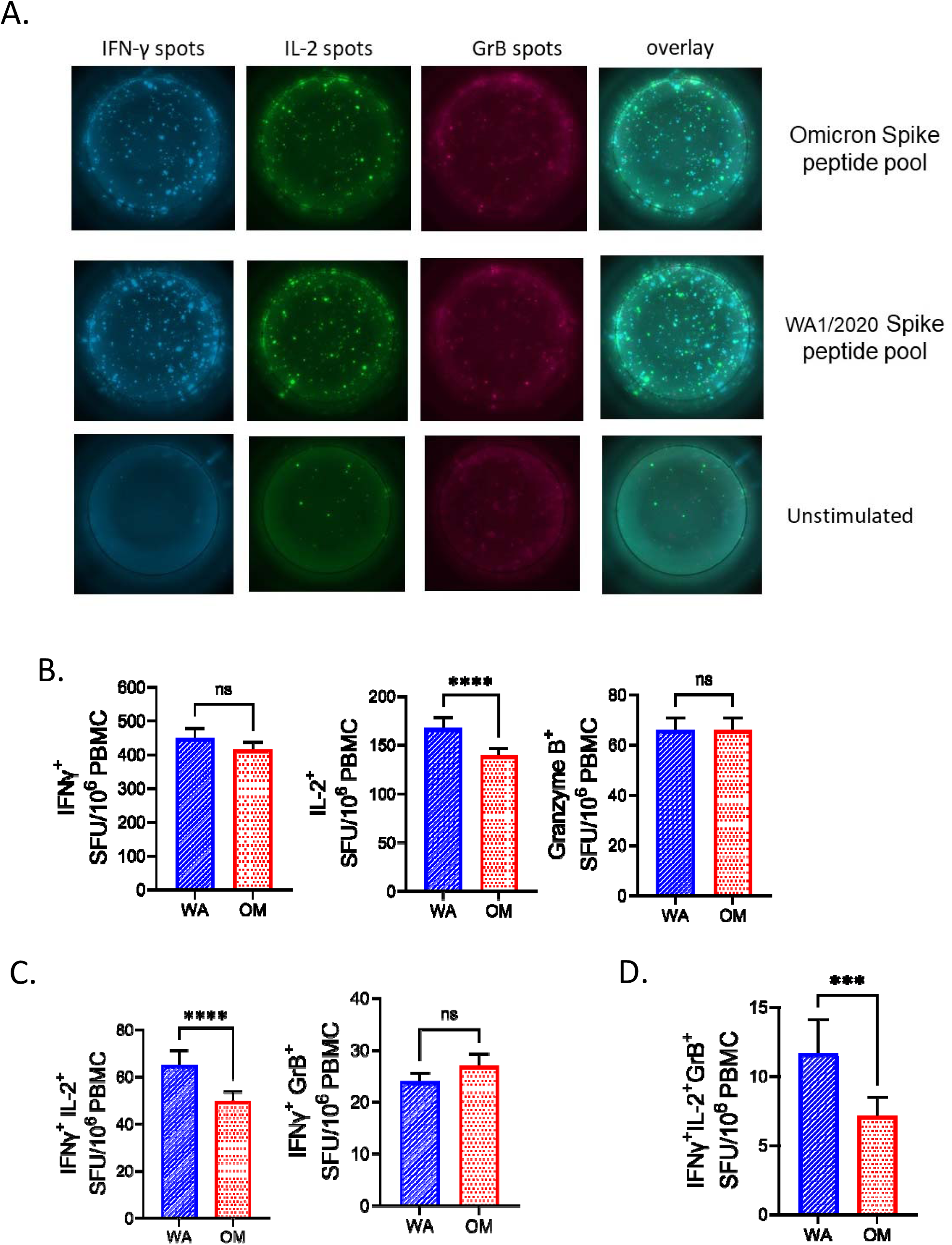
Robust SARS-CoV-2 specific T cells responses in immunized individuals. **A)** 10^6^ PBMCs per well were stimulated with Spike peptide pools from USA-WA1/2020) and B.1.1.529 (Omicron) cultured for 42h in pre-coated IFN-γ, IL-2 and GrB FLUORISpot plates. Both peptide pools induced responses of all three cytokines compared to unstimulated wells. **B)** Ag-specific ELISpot numbers were calculated by subtracting the unstimulated wells of each participant from the peptide stimulated wells. All immunized subjects (post infection and vaccinated) were pooled together (N=326). Omicron peptide pool induced equal IFN-γ and GrB response compared to USA-WA1/2020 but lower IL-2. **C)** Number of polyfunctional IFN-γ+IL-2+ cells but not IFN-γ+GrB+ cells was reduced after stimulation with Omicron spike peptide pool. **D)** Triple positive IFN-γ+IL-2+GrB+ were also reduced following stimulation with Omicron peptides. Data presented as mean ± standard error of the mean. Two-tailed Wilcoxon rank test. For all statistical differences *p<0.05, **p<0.01, ***p<0.001. ****p<0.0001.

We next investigated the differences in T cell responses between the groups of study subjects. When stimulated with the WA1/2020 peptide pool, recipients of two doses of mRNA vaccine had significantly lower numbers of IFN-γ SFU (**Figure 2A**, left panel) than all other groups. A similar trend was measured with the Omicron peptide pool, albeit without reaching statistical significance (**Figure 2A**, right panel). Similarly, number of IL-2 SFU was decreased in the 2X VAX group stimulated with WA1/2020 peptides (**Figure 2B**). Double positive (IFN-γ+IL-2+) and triple positive (IFN-γ+IL-2+GrB+) SFU were also lower in the 2X VAX group (**Figure 2C**). Interestingly, the magnitude of polyfunctional triple positive cells to Omicron was the highest in the 3X VAX group (**Figure 2D**, right panel). Jointly, these results imply that two doses of mRNA vaccine induced sub-maximal T cell immunity, and that at least three doses are required to achieve the same level of antigen specific T cells found in post-infection subjects. To compare the relative preservation of T cell responses to each variant between subjects vaccinated with courses of vaccines produced by different manufacturers (Pfizer and Moderna), we pooled the results from the 2X and 3X vaccinated subjects and divided them into groups based on vaccine manufacturer. We found that while there was no statistical difference in IFN-γ T cell responses between ancestral and Omicron pools within vaccine manufacturer, there was a higher number of IL-2+ SFU detected in vaccines who received Moderna’s vaccine (**Supp. Fig. 2A**). We measured a higher number of IL-2+ SFU in samples from Moderna vaccinated subjects in response to either WA1/2020 or Omicron as compared to their Pfizer vaccinated counterparts (**Supp. Fig. 2B**). The subjects grouped by vaccine manufacturer did not differ in the number of GrB SFU (**Supp. Fig. 2C**). Next, we investigated whether T cell responses were dependent on the age of the subject or time elapsed from vaccination. The 2X VAX subjects were the largest group (N=215), and therefore the most appropriate cohort for this specific analysis. Additionally, analysis of all groups with immune memory demonstrated that IL-2 production was the cytokine most impacted by the mutations in Omicron Spike peptides as compared to WA1/2020 (**Fig. 1**) so we analyzed the impact of age or time from vaccination by IL-2+ SFU. We have detected a minimal, but measurable, negative correlation of IL-2 responses with age (r=-0.1575, p=0.0224) while there was no statistically significant association of IL-2 and time from vaccination (**Figure 3A, B**). There was no impact of age for double positive (IFN-γ+IL-2+) SFU, whereas triple positive (IFN-γ+IL-2+GrB+) SFU were negatively correlated to age of the subject (r=-0.2386, p<0.0001) but not to time-from-vaccination (**Figure 3C**). Overall, changes in function of T cell responses with the age of the participant was minimal. Additionally, given that the 3X vaccinated group was on average 8 years older than 2X VAX (58 vs 50 years average age, respectively, Table 1) and had higher T cell responses than 2X VAX (Figure 2A-2D) we conclude that age is not a major determinant in T cell responses to mRNA vaccines following administration of multiple vaccine doses.

**Figure 2.**
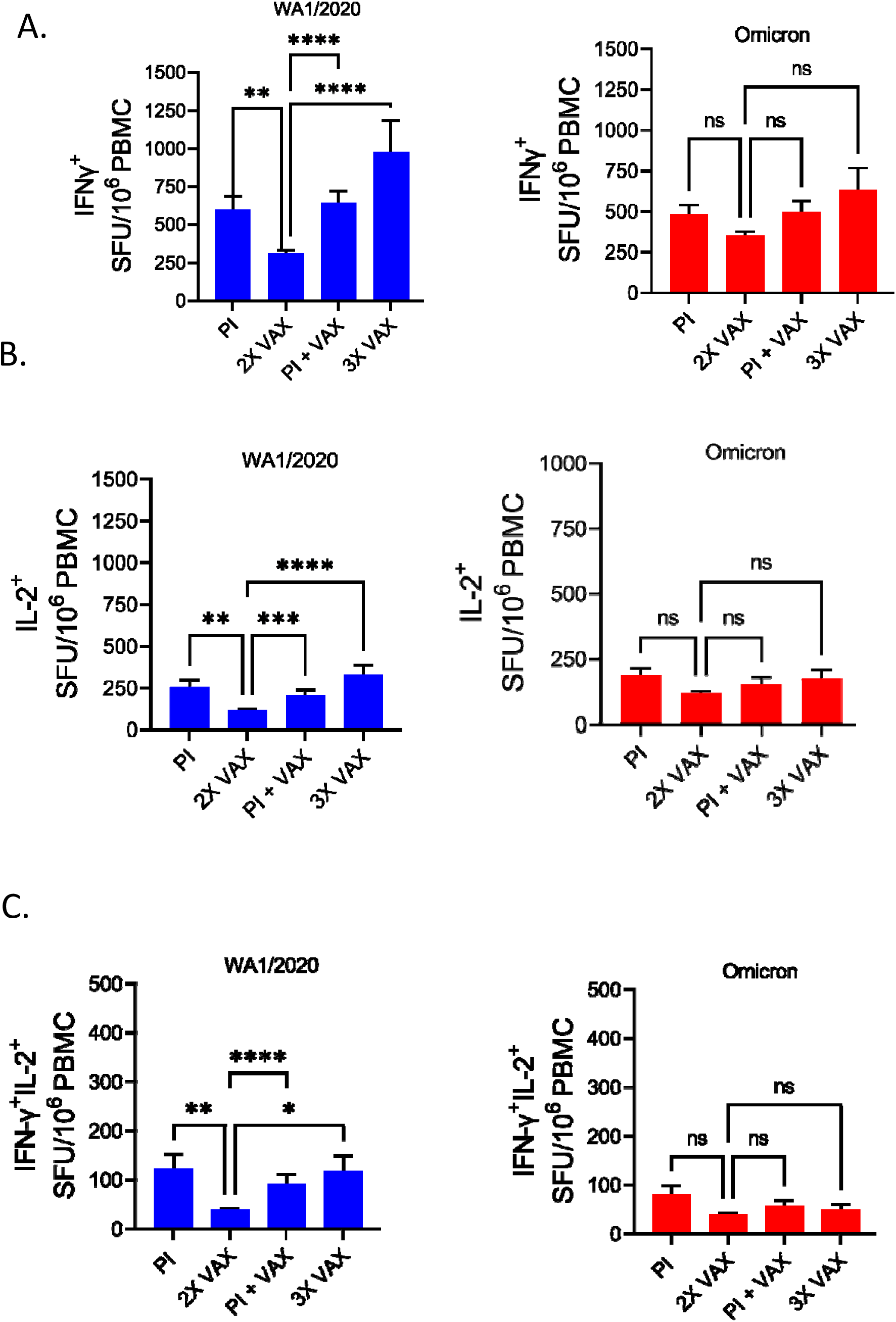
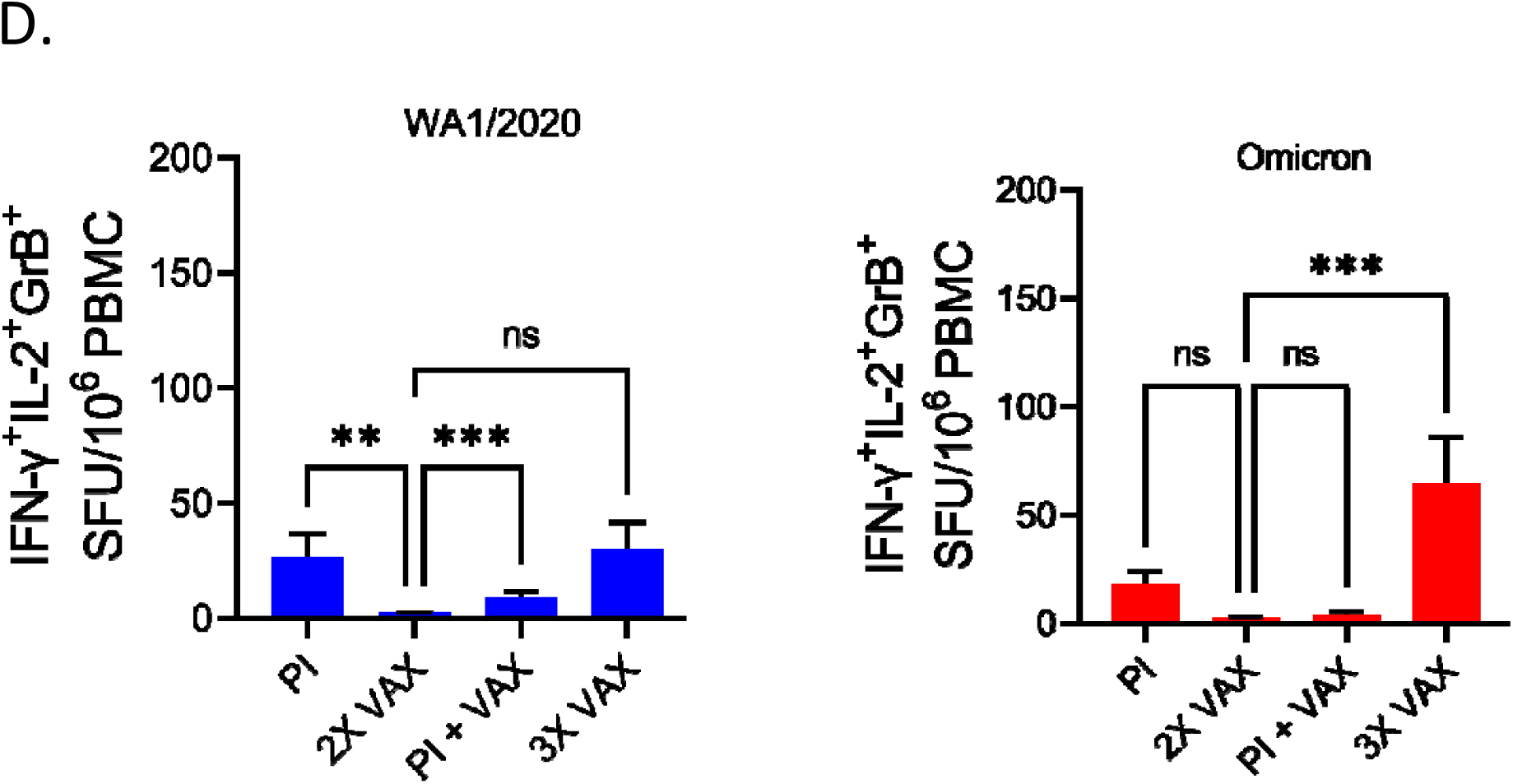
SARS-CoV-2 T cell responses are lower in recipients of two doses of mRNA vaccines. **A)** IFN-γ SFU were reduced in 2X VAX group compared to other groups when their PBMCs were stimulated with USA-WA1/2020 peptide pool (left panel), similar trend was observed with Omicron peptides but without statistical significance. **B)** IL-2 SFU were also reduced in the 2X vax group stimulated with USA-WA1/2020 peptide pool as were **C)** double positive IFN-γ+IL-2+ cells. **D)** number of triple positive IFN-γ+IL-2+GrB+ cells was highest in 3X VAX group. Post infection (PI) n=54, Post infection and vaccinated (PI +VAX) n=39, two dose vaccinated (2X VAX) n=210, three dose vaccinated (3X VAX) n=24.Data presented as mean ± standard error of the mean. Kruskal-Wallis test with Dunn’s correction for multiple comparison

**Figure 3.**
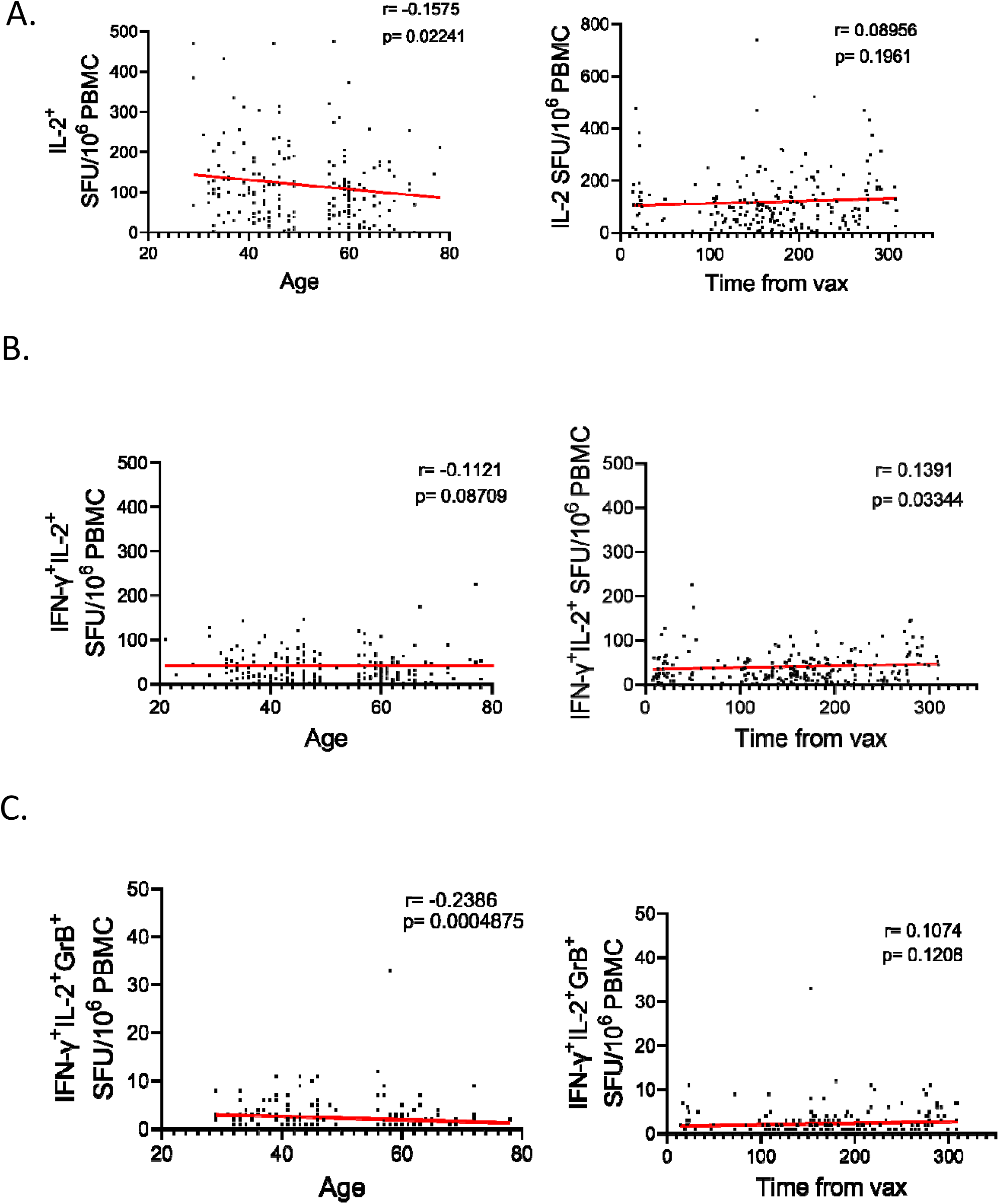
Correlation of the T cell response to age of the subject and time from vaccination in 2X VAX participants. **A)** IL-2 response to Omicron peptide pool was weakly negatively correlated to age (r=-0.1575. p=0.02241) but not to time from vaccination (p=0.1961). *B)* no correlation with age was observed for double positive IFN-γ+IL-2+ cells (p=0.087). **C)** triple positive IFN-γ+IL-2+GrB+ were weakly negatively correlated with age (r=-0.2386, p<0,001). n=210, nonparametric Spearman correlation, two tailed p-values. Data presented as individual values.

To further interrogate the phenotype of T cells responding to either WA1/2020 or Omicron we performed flow cytometry (FCM) on approximately half of the 2X VAX subject samples (randomly selected, n=96). Expression of costimulatory molecules CD137 (4-IBB) and OX-40 is the most common method for enumerating antigen (ag) specific T cells by flow cytometry^18^. Again, as with the FLUORISpot assay, Spike peptide pool induced a robust increase in CD137 (4-IBB) and OX-40 activated T cells (CD137+OX-40+) compared to stimulated wells (**Suppl. Figure 3A**, representative FCM in **Figure 4A**). As previously reported^19^, we have detected more CD4 helper T cells than cytotoxic CD8 T cells upregulating CD137 (4-IBB) and OX-40 in response to spike peptide pools (**Figure 4B**). This was further reinforced by higher correlation of the FLUORISpot response with flow cytometric CD4+CD137+OX-40+ cells than CD8+CD137+OX-40+ for both IFN-γ (**Figure 4C**) and IL-2 (**Figure 4D**). We did detect a slight decrease in the number of ag-specific CD4 and CD8 T cells in response to Omicron peptides vs. WA1/2020. However, the difference was not statistically significant (**Figure 4B**). We also analyzed the naïve and memory phenotype of circulating T cells for possible correlation to ag-specific T cell responses. We did not find any association of the naïve circulating T cell phenotype (CCR7+CD45RA+CD28+CD95-) or the advanced differentiation/senescence-related marker CD57 with antigen specific response (**Supplemental Figure 3B and C**.). Given that CD4+ helper T cells represent the bulk of the Spike peptide response we have additionally investigated the expression of Th polarizing cytokines IL-17a and IL-4 in the 2X VAX group. We found that both IL-17a (**Figure 4E**) and IL-4 (**Figure 4F**) were reduced in response to Omicron peptides, which could be interpreted as a positive finding, given that Th17 and Th2 responses would be expected to be counterproductive against viral infection.

**Figure 4.**
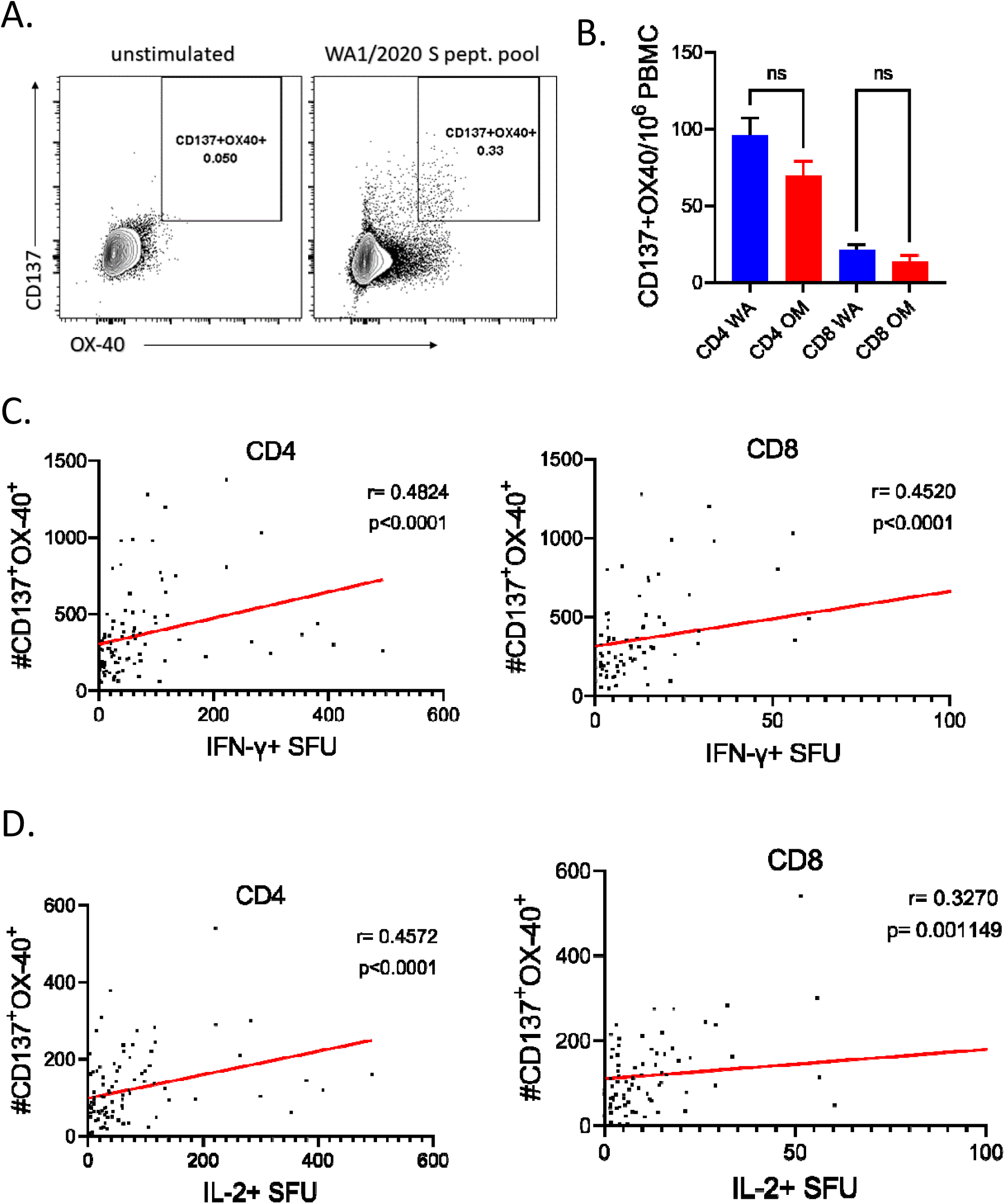
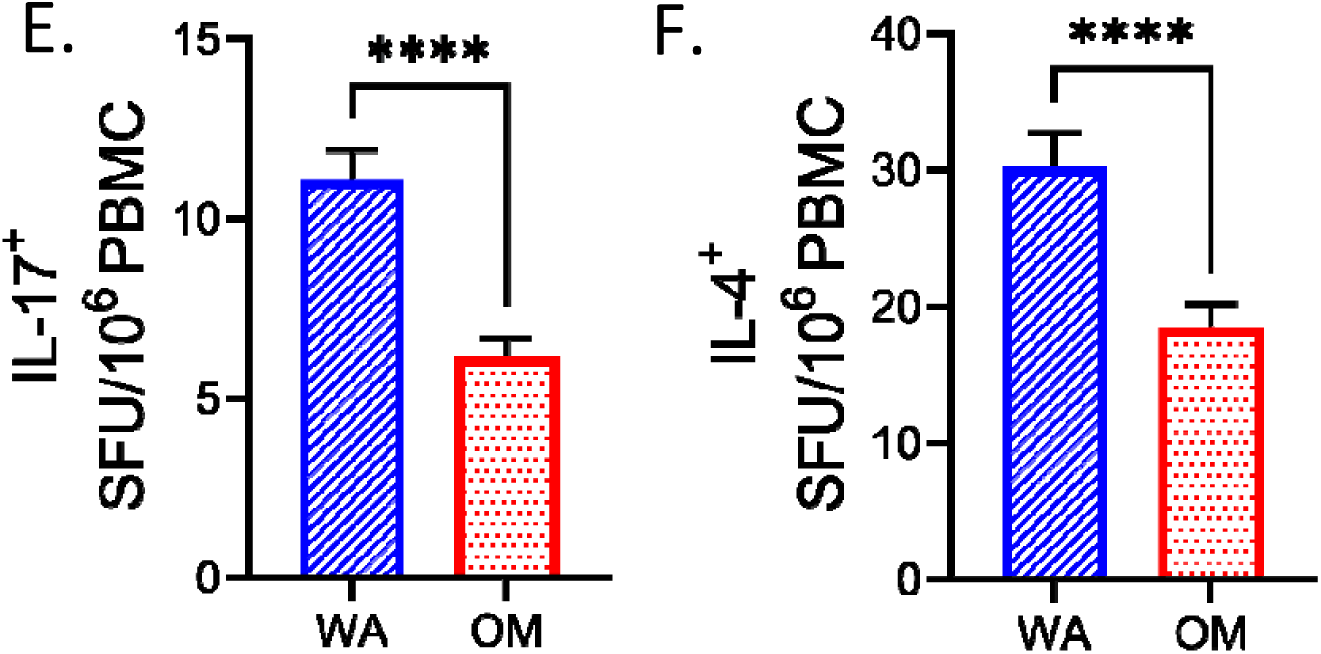
Spike peptide pools induce a greater helper (CD4+) T cell response than cytotoxic (CD8+) T cell response. **A)** representative flow cytometric plot of Spike peptide induction of expression of costimulatory molecules CD137 and OX-40 on T cells. **B**. Number of CD4+ cells expressing CD137 and OX-40 was higher than for CD8 T cells. N=96, data presented as mean ± standard error of the mean. Kruskal-Wallis test with Dunn’s correction for multiple comparisons **C**. Correlation between IFN-γ SFU or **D**. IL-2 SFU measured by FLUORISpot and expression of CD137 and OX-40 measured by flow cytometry was higher for CD4 than CD8 T cells. N=96, data presented as individual values, nonparametric Spearman correlation, two tailed p-values. **E)** IL-17a and **F)** IL-4 were reduced in response to Omicron peptides in subjects which received two doses of mRNA vaccines. n=96, data presented as mean ± standard error of the mean. Two-tailed Wilcoxon rank test. For all statistical differences *p<0.05, **p<0.01, ***p<0.001. ****p<0.0001.

Overall, our findings highlight resilience of T cell responses generated in response to a pre-Omicron infection and/or to Wuhan-derived spike protein-based vaccines in the face of the B1.1.529 Omicron variant, particularly as measured by the IFN-γ production, which was by far the largest component of the antiviral T cell response. It is tempting to speculate that this finding may in part explain resistance against severe forms of disease and death in breakthrough Omicron cases. We did measure decreased IL-2 and IL-2 plus other cytokines (polyfunctional) responses, however these were generally reduced by less than 20-30%, and the extent of reduction depended on the exposure/vaccination, being less reduced, if at all, in three-doses vaccine recipients. Stratified analysis of subjects immunized by vaccines or prior infection clearly revealed that 2X VAX participants exhibited inferior responses to both Omicron and the ancestral strain relative to infected, infected and vaccinated, and in particular, 3X VAX participants, that trended, or were significantly higher, by most measures of immunity. Therefore, our results stress the need for a three-dose vaccine regimen to achieve robust T cell immunity.

## Data Availability

All data produced in the present work are contained in the manuscript.

## Acknowledgements

We would like to thank the personnel of UArizona Health Sciences Biobank for expert processing and storage of human blood samples. We would like to thank our CDC colleagues Drs Natalie J. Thornburg, Mark G. Thompson and Julie Mayo Lamberte for logistical help, input into study design and comments on the manuscript.

## Conflict Statement

Dr Nikolich is receiving research support, as well as honoraria as co-Chair of the Scientific Advisory Board for Young Blood Institute, Inc.,(YBI), a non-profit company with the goal to explore the efficacy of use of human plasma and other blood products to delay adverse effects of aging and/or mitigate chronic diseases. YBI had no impact on the design, execution or conclusions of the present study or on the contents of this manuscript.

## Notes

Supported in part by the USPHS award R37 AG020719 and the Bowman Professorship in Medical Sciences to J.N-Z. and the CDC HEROES project award 75D30120C08379 to J.L.B.

### Author Declarations

This study was approved by the University of Arizona IRB (protocol #2102460536 and #1410545697) and the Oregon Health and Science University IRB (protocol #00003007, pre-pandemic controls) and was conducted in accordance with all federal state and local regulations and guidelines.

## References

1. Karim, S. S. A. & Karim, Q. A. Omicron SARS-CoV-2 variant: a new chapter in the COVID-19 pandemic. Lancet 398, 2126–2128 (2021).

2. CDC. Coronavirus Disease 2019 (COVID-19). Centers for Disease Control and Prevention https://www.cdc.gov/coronavirus/2019-ncov/science/science-briefs/scientific-brief-omicron-variant.html (2020).

3. Investigation of SARS-CoV-2 variants: technical briefings. GOV.UK https://www.gov.uk/government/publications/investigation-of-sars-cov-2-variants-technical-briefings.

4. Statement – Update on COVID-19: Omicron is gaining ground: Protect, prevent, prepare. https://www.euro.who.int/en/media-centre/sections/statements/2021/statement-update-on-covid-19-omicron-is-gaining-ground-protect,-prevent,-prepare..

5. CDC. COVID Data Tracker. Centers for Disease Control and Prevention https://covid.cdc.gov/covid-data-tracker (2020).

6. HKUMed finds Omicron SARS-CoV-2 can infect faster and better than Delta in human bronchus but with less severe infection in lung. https://www.med.hku.hk/en/news/press/20211215-omicron-sars-cov-2-infection.

7. Lupala, C. S., Ye, Y., Chen, H., Su, X. & Liu, H. Biochemical and Biophysical Research Communications Mutations on RBD of SARS-CoV-2 Omicron variant result in stronger binding to human ACE2 receptor. Biochem. Biophys. Res. Commun. 590, 34–41 (2022).

8. Wilhelm, A. et al. Reduced Neutralization of SARS-CoV-2 Omicron Variant by Vaccine Sera and monoclonal Abstract_: (2021).

9. Rössler, A., Riepler, L., Bante, D., Laer, D. von & Kimpel, J. SARS-CoV-2 B.1.1.529 variant (Omicron) evades neutralization by sera from vaccinated and convalescent individuals. medRxiv 2021.12.08.21267491 (2021) doi:10.1101/2021.12.08.21267491.

10. Maslo, C. et al. Characteristics and Outcomes of Hospitalized Patients in South Africa During the COVID-19 Omicron Wave Compared With Previous Waves. JAMA (2021) doi:10.1001/jama.2021.24868.

11. Abdullah, F. et al. Decreased severity of disease during the first global omicron variant covid-19 outbreak in a large hospital in tshwane, south africa. Int. J. Infect. Dis. (2021) doi:10.1016/j.ijid.2021.12.357.

12. McMahan, K. et al. Correlates of protection against SARS-CoV-2 in rhesus macaques. Nature 590, 630–634 (2021).

13. Rydyznski Moderbacher, C. et al. Antigen-Specific Adaptive Immunity to SARS-CoV-2 in Acute COVID-19 and Associations with Age and Disease Severity. Cell 183, 996-1012.e19 (2020).

14. Liao, M. et al. Single-cell landscape of bronchoalveolar immune cells in patients with COVID-19. Nat. Med. 26, 842–844 (2020).

15. Jergovič, M. et al. Competent immune responses to SARS-CoV-2 variants in older adults following mRNA vaccination. bioRxiv 2021.07.22.453287 (2021) doi:10.1101/2021.07.22.453287.

16. Tarke, A. et al. Article Impact of SARS-CoV-2 variants on the total CD4 + and CD8 + T cell reactivity in infected or vaccinated individuals ll ll Impact of SARS-CoV-2 variants on the total CD4 + and CD8 + T cell reactivity in infected or vaccinated individuals. Cell Reports Med. 2, 100355 (2021).

17. Lutrick, K. et al. COVID-19 Infection, Reinfection, and Vaccine Effectiveness in Arizona Frontline and Essential Workers_: Protocol for a Longitudinal Cohort Study Corresponding Author_: 10, 1–13 (2021).

18. Grifoni, A. et al. Article Targets of T Cell Responses to SARS-CoV-2 Coronavirus in Humans with COVID-19 Disease and Unexposed Individuals ll Article Targets of T Cell Responses to SARS-CoV-2 Coronavirus in Humans with COVID-19 Disease and Unexposed Individuals. Cell 181, 1489-1501.e15 (2020).

19. Guerrera, G. et al. BNT162b2 vaccination induces durable SARS-CoV-2 – specific T cells with a stem cell memory phenotype. 1–13 (2021).

