## Supplemental Figures for "Resilient T cell responses to B.1.1.529 (Omicron) SARS-CoV-2 variant"

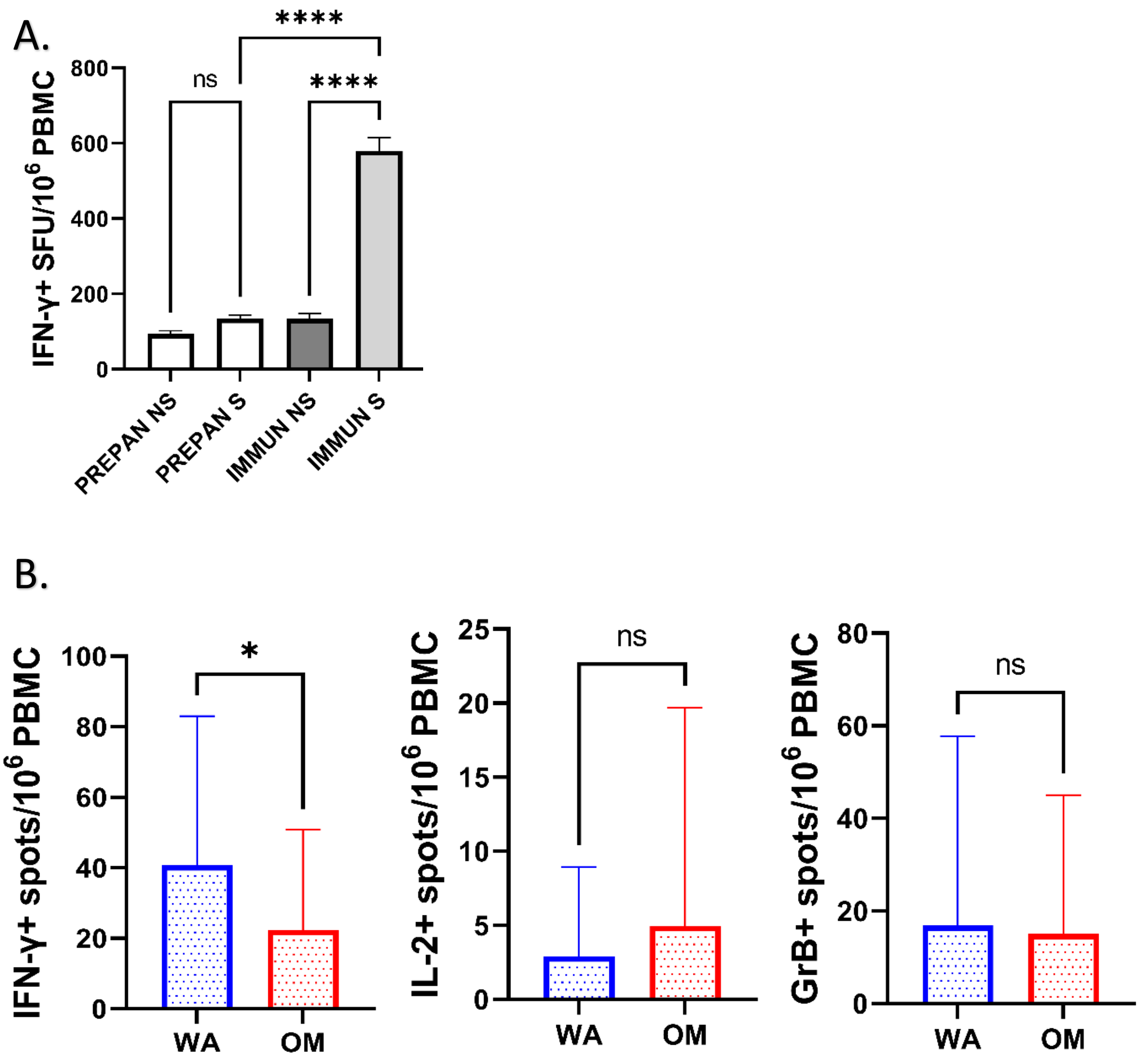


**Supplemental Figure 1. A)** $10^6$ PBMCs per well were cultured without peptide stimulation (NS) or stimulated with Spike peptide pools from USA-WA1/2020 for 42h, IFN-γ response was measured by FLUORISpot. Prepandemic samples N=32, immunized samples N=326. Data presented as mean ± standard error of the mean. Kruskal-Wallis test with Dunn's correction for multiple comparisons. **B).** Prepandemic samples showed higher IFN-γ responses, but not IL-2 and GrB in response to USA-WA1/2020 peptide pool compared to Omicron. N=32. Data presented as mean ± standard error of the mean. Two-tailed Wilcoxon rank test. For all statistical differences *p<0.05, **p<0.01, ***p<0.001. ****p<0.0001.

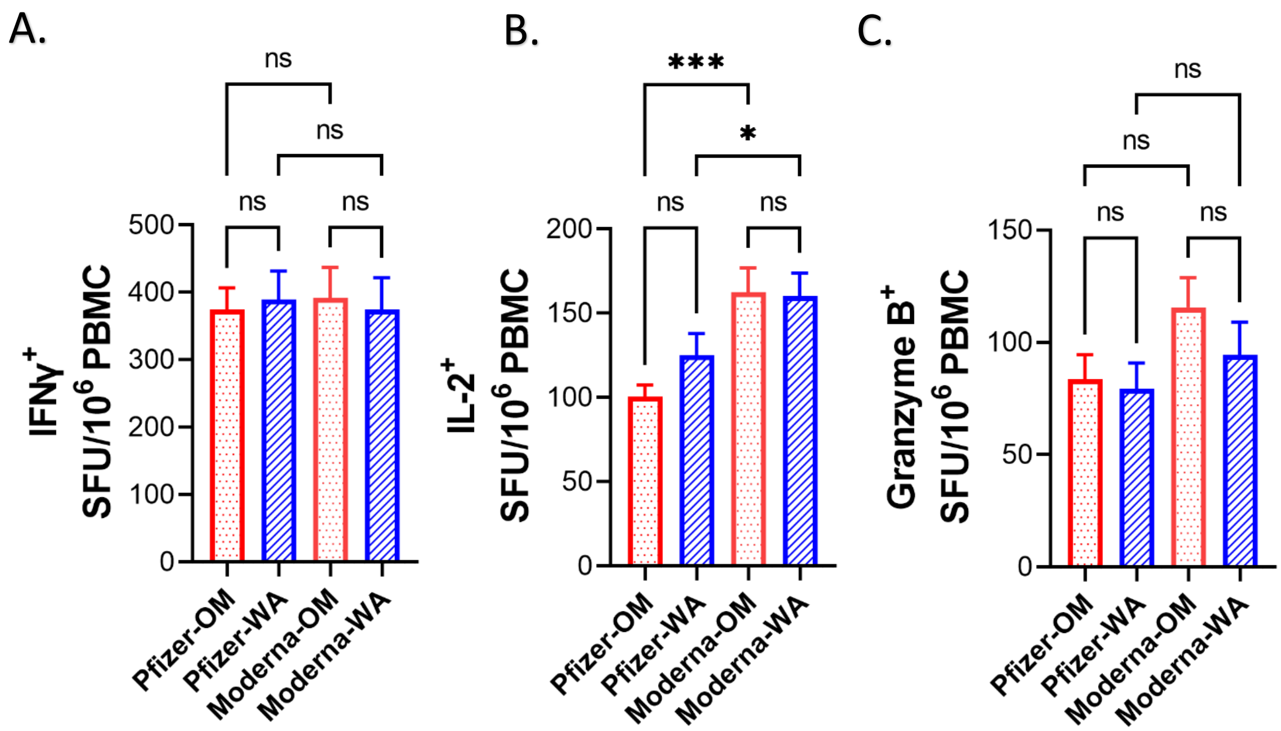


**Supplemental Figure 2. Reduced IL-2 responses in recipients of Pfizer mRNA SARS-Cov-2 vaccines A)** No difference in number of IFN-γ SFU between recipeients of Pfizer and Moderna manufactured mRNA vaccines. **B)** IL-2 SFU were reduce in recipient of Pfizer's mRNA vaccines in response to both USA-WA1/2020 and Omicron peptide pool. C) No difference in number of GrB SFU between group. Pfizer n=139, Moderna n=93. Data presented as mean ± standard error of the mean. Kruskal-Wallis test with Dunn's correction for multiple comparisons

| Marker | Fluorochrome | Clone |
|---|---|---|
| CD16 | PerCPCY5.5 | 3G8 |
| CD3 | AF488 | UCHT1 |
| CD4 | SB550 | SK3 |
| CD8 | AF700 | SK1 |
| CD45RA | BV570 | HI100 |
| CD28 | PEDazzle994 | CD28.2 |
| CD95 | BV421 | DX2 |
| CCR7 | PECy7 | G043H7 |
| OX-40 | PE | Ber-ACT35 (ACT35) |
| CD137 | APC | 4-1BB |
| CD57 | BV605 | QA17A04 |

**Supplemental Table 1. Antibodies used for flow cytometric staining**

A.

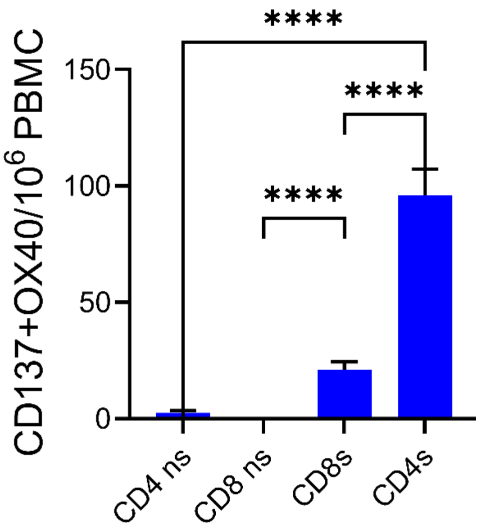

****
****
****
CD137+OX40/10^6 PBMC
150
100
50
0
CD4 ns
CD8 ns
CD8s
CD4s


B.

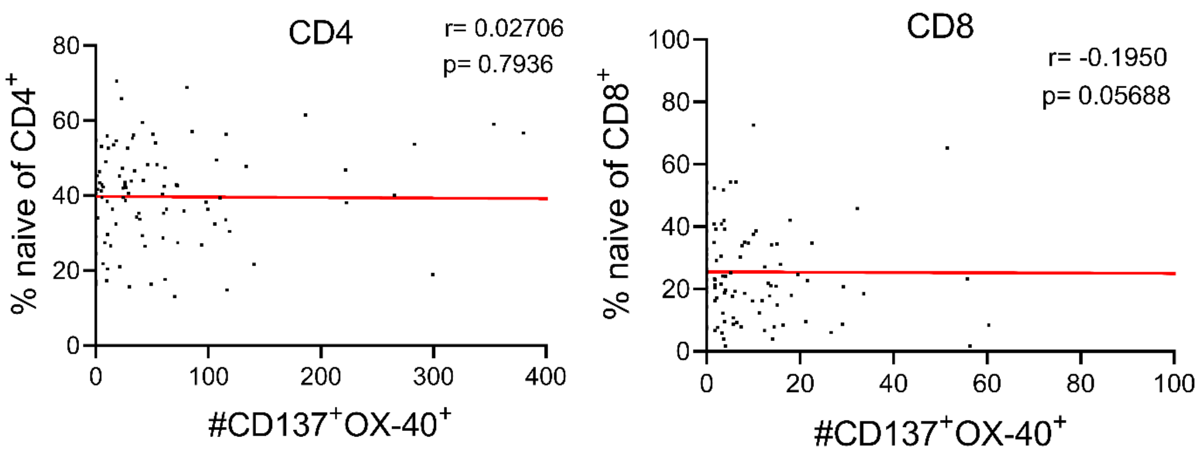

CD4
r= 0.02706
p= 0.7936
% naive of CD4+
#CD137+OX-40+
CD8
r= -0.1950
p= 0.05688
% naive of CD8+
#CD137+OX-40+


C.

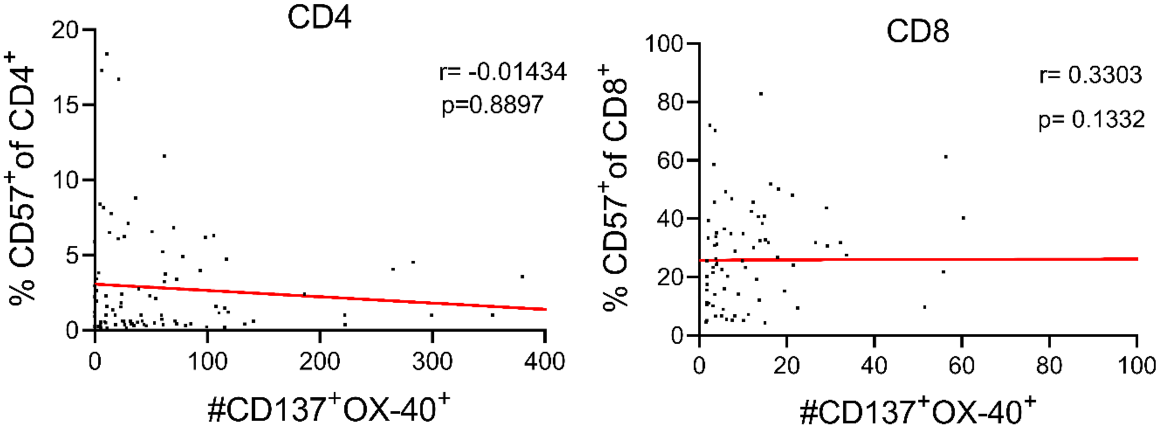

CD4
r= -0.01434
p=0.8897
% CD57+of CD4+
#CD137+OX-40+
CD8
r= 0.3303
p= 0.1332
% CD57+of CD8+
#CD137+OX-40+

**Supplemental Figure 3. A)** Spike peptide pool induces expression of CD137 and OX-40 on T cells, n=96. Data presented as mean ± standard error of the mean. Kruskal-Wallis test with Dunn's correction for multiple comparisons **B)** No correlation with antigen specific (CD137+OX-40+) cells and naïve phenotype of T cells or **C)** senescence marker CD57, n=96, nonparametric Spearman correlation, two tailed p-values. Data presented as individual values.
